# Measurement of motivation states for physical activity and sedentary behavior: Development and validation of the CRAVE scale

**DOI:** 10.1101/2020.08.31.20184945

**Authors:** Matthew A. Stults-Kolehmainen, Miguel Blacutt, Nia Fogelman, Todd A. Gilson, Philip R. Stanforth, Amanda L. Divin, John B. Bartholomew, Alberto Filgueiras, Paul C. McKee, Garrett I. Ash, Joseph T. Ciccolo, Line Brotnow Decker, Susannah L. Williamson, Rajita Sinha

## Abstract

Physical activity, and likely the motivation for it, varies throughout the day. The aim of this investigation was to create a short assessment (CRAVE) to measure motivation states (wants, desires, urges) for physical activity and sedentary behaviors. Five studies were conducted to develop and evaluate the construct validity and reliability of the scale, with 1,035 participants completing the scale a total of 1,697 times. In Study 1, 402 university students completed a questionnaire inquiring about the want or desire to perform behaviors “at the present moment (right now)”. Items related to physical activity (e.g., “move my body”) and sedentary behaviors (e.g., “do nothing active”). An exploratory structural equation model (ESEM) revealed that 10 items should be retained, loading onto two factors (5 each for Move and Rest). In Study 2, an independent sample (n= 444) confirmed these results and found that Move and Rest desires were associated with stage-of-change for exercise behavior. In Study 3, 127 community-residing participants completed the CRAVE at 6-month intervals over two years-two times each session. Across-session interclass correlations (ICC) for Move (ICC = .72-.95) and Rest (ICC = .69-.88) were higher than when when they were measured across 24-months (Move: ICC = .53; Rest: ICC = .49), indicating wants/desires have state-like qualities. In Study 4, a maximal treadmill test was completed by 21 university students. The CRAVE was completed immediately pre and post. Move desires decreased 26% and Rest increased 74%. Changes in Move and Rest desires were moderately associated with changes in perceived physical fatigue and energy. In Study 5, 41 university students sat quietly during a 50-minute lecture. They completed the CRAVE at 3 time points. Move increased 19.6% and Rest decreased 16.7%. Small correlations were detected between Move with perceived energy and tiredness, but not calmness or tension. In conclusion, the CRAVE scale has good psychometric properties. Data also support tenets of the WANT model of motivation states for movement and rest (Stults-Kolehmainen et al., 2020). Future studies need to explore how desires to move/rest relate to dynamic changes in physical activity and sedentarism.

## Introduction

Conditions such as obesity, cardiovascular disease (CVD) and diabetes are all related to a lack of muscular movement and minimal energy expenditure (EE) (1). EE, however, is affected by distinct *behaviors* falling in two dichotomous categories: physical activity (i.e., occupational activity, active transit, exercise) and sedentary behavior (e.g., sitting, watching TV, sleep). These are ostensibly separate but not mutually exclusive constructs, which have independent and interacting effects on health (2-4). Recent evidence indicates that humans display substantial variability in engaging in active and sedentary behaviors. There is also asymmetry in these behaviors. For instance, some individuals achieve high levels of activity, but also high levels of sedentarism (5). Given the distinction between physical activity (PA) and inactivity, growing importance is being placed on understanding the various genetic, neurological, environmental and psychological antecedents or determinants of these two behaviors (6, 7). Motivation appears to be a key intermediary of physically active behaviors. Unfortunately, there is a gap in our current understanding of how the environment and the brain interact to actually motivate movement and sedentary behaviors, particularly on a *moment to moment basis*, which suggests that additional mechanisms may be responsible for their linkage (6).

Research in the area of motivational processes greatly lags behind work in the areas of cognition and emotion, but newer models are reviving older perspectives and synthesizing this with data that suggests that motivated behavior starts with *motivation states*, such as *desires* and *urges* (8-17). Several dual-process models of physical activity have recently emerged-the Affective Reflective Theory of Physical Inactivity and Exercise (ART) (18), the model from Conroy and Berry (19), the Affect and Health Behavior Framework (AHBF) (20) and the later Integrated Framework (AHBF-IF) (15). All these models view physical activity and sedentary outcomes as the consequence of interactions between two systems, one responsible for affective processing and one responsible for intentional planning, goal setting and other cognitive processes. Brand and Ekkekakis (18) incorporate Lewin’s (21) ideas of driving and restraining forces, with the interplay resulting in tensions, which he sees as response to a need, want or some other stimulus, resulting in an “intention or desire” to carry out a specific task (22). In ART, the affective system operates with automatic processes of stimulus conditioning and hedonic value assessment resulting in an *action impulse*, which is the antecedent to physically active behavior. They also note that “…core a□ective valence may have a direct, immediate impact on behavior through behavioral urges” (18). The Affect and Health Behavior Framework (AHBF) model (20) and its successor, the Integrated Framework, (AHBF-IF) (15), specifies that the faster, affective system operates on behavior through two intermediaries: automatic motivation (i.e., “wanting”, “dread”) and affectively charged motivational states (ACMS) (i.e., “craving & desire”, “fear”) (23), which interact with goals and intentions to influence physically active behaviors. A total lack of desire is a hallmark of amotivation as outlined in Self-Determination Theory (24-26). Finally, in ancient philosophy, Aristotle concluded, “It is manifest, therefore, that what is called desire is the sort of faculty in the [mind] which initiates movement” (27-30). Consequently, there appears to be widespread theoretical support for the idea that motivation states for movement, such as wants or urges, propel physical activity.

Given the recent development of most of these theories, however, one needs to cast a wider net across the literature to find empirical evidence of wants and urges to move the physical body. A preponderance of information about desires for movement exists in clinical and psychiatric literatures. Urges to move, especially at night, are a defining characteristic of Restless Leg Syndrome, which may affect other parts of the body as well (31-33). Exercise dependence/addiction is a “craving for leisure-time physical activity, resulting in uncontrollable excessive exercise behavior” and showing a variety of physical and psychological symptoms (34). Long-term use of anti-psychotic medication can result in psychogenic movement disorders, such as tardive dyskinesia and akathisia, the latter of which results in a pressing need to constantly fidget and move about (35). Dopamine receptor-blocking agents can also induce tardive dyskinesia, which results in an inner urge to move that manifests in chorea, dystonia, akathisia, tics and tremors (36). Aside from these clinical presentations, desires to move may manifest in response to environmental conditions, like upbeat rhythmic music. As an example, *groove* is a state of desiring to move the body in response to a rhythm or harmony (37-41). Other researchers have identified desires as being specific to fitness, for instance, a desire to gain muscular strength (42) or even to generally engage in sport (12, 43). Even animals clearly demonstrate urges to move when deprived of it, in what is known as *appetence* for muscular motion (9, 44). Interestingly, until recently, none of this literature was examined as a unified body of knowledge.

How feelings of urges, wants, desires and cravings for movement, as well as sedentary behavior, relate and differ is addressed by the WANT (Wants and Aversion for Neuromuscular Tasks) model (17). In developing this heuristic, Stults-Kolehmainen and colleagues define wants and urges to move as “…affectively-charged motivation states and associated feelings that signal a pressing need to approach or avoid a state of muscular movement (or, conversely a state of rest)” (17). Desires or urges to move might be due to the natural drive to move (44-46) which results in tension and is only satisfied when released with movement, which is considered negative reinforcement (17, 47). In this model, wants and urges for movement and rest are considered separate factors relating orthogonally on two different axes/continua as opposed to opposite poles of the same continuum, similar to a circumplex configuration (48). The WANT model specifies that wants/desires to move and rest typify psychological states that are highly transient and change with varying conditions, like physically taxing conditions (e.g., a hard workout), stressful circumstances and conflicting situations when one might be both very tired and also energetic (e.g., having just won a competition but also wanting to celebrate) (49). Where they cross at their 0 points, it is theorized that humans inhabit a deactivated states, perhaps similar to sleep or meditation (49). Alternatively, the high marks in each direction denote active approach (e.g., in the midst of a competition) of movement or rest (e.g., crawling into bed). The model also specifies times when there is active avoidance of movement (“diswants”), which might be most obvious in instances of fatigue, illness and injury (50), chronic pain and kinesiophobia (51). Unfortunately, this model has not been tested rigorously to understand the strength of the association between desires/urges and physically active behaviors (e.g., exercise, sitting time).

At this juncture, intersecting lines of research strongly suggest that humans possess wants/desires for movement and rest behaviors (8, 15, 17, 35, 38, 52). However, a review study in this special issue reports that “little to no research has been conducted to directly measure craving/desire for PA” (53). Most importantly, scientific inquiry to strengthen our understanding of these desires is currently impeded by a lack of validated instrumentation (54, 55). Only surveys with only 1-3 items measuring desire or urge for muscular movement-related concepts exist, such as of state motivation for specific exercise tasks (56), exercise -in general (57), physical activity (58) or other bodily movement, vaguely defined (59, 60). Simple measures have also been developed to assess desire to move specifically in response to music (39-41) or desires for fitness (42). Some scales refer to wants/desires as a stable construct or trait rather than a state (61). Some ask respondents to look back retrospectively on their desires and urges rather than in the current moment (60). All of these measures lack validation with the exception of a measure to urge to move in the context of akathisia (62). On the other hand, substantial development of instruments to assess desire, wanting and craving for reward substances, foods and behaviors (63-66) and neurobiological substrates associated with desire and craving states (67) has occurred.

The primary aim of the current investigation, therefore, was to create and validate an instrument to measure subjective ratings of desire or want, affectively-charged motivation states (ACMS), for physically active behaviors. In accordance with the WANT model (17), we also aimed to also assess desires/wants for sedentary behaviors. Our objective was to create a measure of desires/wants that will have acceptable reliability, construct validity, and more specifically, convergent validity. A second aim of this investigation was to test premises of the WANT model, which can be accomplished with the same data and analyses. Based on this framework, we generally hypothesized that: 1) humans have a quantifiable/measurable motivation to move that may be perceived as a desire or want (or in higher levels-urge, craving), 2) desires/wants for movement/physical activity are separable from desires/wants to rest or be sedentary, 3) desires/wants are transitory and possess state-like qualities (i.e., pertaining to processes happening in the moment), 4) motivational states for movement and rest change with relevant provision or avoidance of physical stimuli, such as maximal exercise and a period of rest, and 5) desire to move/rest will be related to, but distinct from, psychosomatic sensations, such as perceptions of physical energy, fatigue, tension and calmness. How these hypotheses are operationalized and link to specific studies 1-5 is described below for each study.

The experimental approach for this investigation was rooted in the classic article on scale development from Clark and Watson (68, 69), who assert that construct validity can only be established through a series of studies that involves many observations and may include: examining a factor structure, correlations with other measures, changes over time, changes across an experimental manipulation and differentiations between groups. Consequently, to develop an instrument and test hypotheses related to the WANT model, five total studies were conducted with 4 college-aged populations and 1 community-dwelling sample to psychometrically validate a questionnaire focused on desires/wants for movement/physical activity and rest/sedentarism. In Studies 1 and 2, the scale was developed and distributed and factor analyses were conducted. Study 3 followed participants multiple times over a two-year period with the same scale. Study 4 monitored changes in affectively-charged motivation states before and after a treadmill test. Finally, Study 5 administered the questionnaire 3 times over a 50-minute lecture period.

## Study 1

### Introduction

No validated instrument exists to measure desires/wants for physical activity and sedentary behaviors, despite wide interest from across several literatures, summarized by Stults-Kolehmainen et al. (17), a number of non-validated scales in publication (cited above) and calls for a validated instrument to be created (54). The primary purposes of study 1 were to, a) construct and b) begin the first steps of validation of a scale for affectively-charged motivation states (ACSM; desires/wants) to move and rest. The secondary purpose, as per above, was to test provisions of the WANT model; the first three major hypotheses from above were addressed. For the first hypothesis, it is predicted that participants will rate themselves as having desires/wants for movement and rest, indicated by score means and variability > 0 (based on t-tests). It is predicted that, for hypothesis 2, an exploratory structural equation model (ESEM) will determine that desires/wants for movement are separate from those for rest (i.e., 2 factors will emerge). However, it could also be argued that additional desire/want factors other than muscular movement and rest may emerge (e.g., a desire or want for *stillness*, an outcome of meditation) (49, 70-72). For the third hypothesis, we predict that desires/wants reported in the current moment (“right now”) will significantly vary from those surveyed retrospectively “over the past week” (60).

### Methods

#### Participants

A total of 402 college students from a public Midwestern university participated in the first study (*M*_age_ = 20.9 years, *SD* = 3.2). Self-reported data revealed that 61.6% of participants were male and 64.9% Caucasian, 17.7% African American, 6% Hispanic/Latino(a), 5% Asian American, 4.7% multiple ethnicities, and 1.5% other/not listed. The academic rank of students was evenly distributed and ranged from a low of 23.1% for sophomores to 26.6% for seniors. Finally, most students (95%) were not athletes on varsity teams sponsored by the university. All individuals signed an informed consent. The study was approved by the Institutional Review Board at Northern Illinois University, in accordance with the Declaration of Helsinki, protocol # HS13-0035.

#### Instrumentation

In addition to a demographic questionnaire, participants completed the Cravings for Rest and Volitional Energy Expenditure (CRAVE) consisting of 30 questions related to the wants/desires or urges individuals had to perform various behaviors or activities. The questionnaire was divided into two identical 15-question parts; the first of which asked participants to respond based on their want/desire to behave in the manners listed *over the past week* (WEEK), while the second was focused on individuals’ want/desire to perform the behaviors *at this very moment / Right now* (NOW) (60, 73-75). Items for each part of the questionnaire were generated by three researchers (MSK, TG and RS) who have research experience in the areas of physical activity, kinesiology, and psychology of addictive behaviors. A process of trimming resulted in 15 acceptable items, seven items related to being physically active (e.g., want/desire to burn some calories; be physically active; exert my muscles; move around etc.) (60), while eight items were sedentary behaviors (e.g., want/desire to just sit down, do nothing active, rest my body, etc.). These items were then checked for face validity by four expert scholars (noted for being experts in the field of exercise psychology) from four different institutions who all endorsed the items (JB, JC and two in the acknowledgements section). Participants responded to each item on an 11-point Likert Scale ranging from 0 = not at all, to 10 = more than ever (36). A full description of the scale development is provided in Supplement 1.

#### Procedure: data collection

After receiving Institutional Review Board approval, two researchers from the byline reached out to professors at the selected university who instructed classes of more than 50 students soliciting participation by briefly describing the nature of the study and the approximately 10-minute time commitment required. Seven professors agreed to the request and upon the prearranged date/time one author traveled to each class for data collection. When addressing the students, the researcher explained the purpose/goals of the study, participants’ rights if they elected to participate, and that all data were anonymous. Students were then allowed time to ask any questions or opt out of participation (no student declined participation). Questionnaires were distributed and collected by the researcher and they were completed at the beginning of each class after written consent.

#### Data Analysis

Data were double entered, cleaned for extreme responders (e.g. those who answered all top or all bottom on all items), and screened for univariate outliers in SPSS version 26 (Armonk, NY, IBM Corporation). No cases were eliminated due to missing data. As latent factor detection is dependent on covariances (76), correlations amongst the items were assessed. Since the items were repeated at two time points (“Right Now” and “Past Week”), correlations were conducted at each timepoint separately. Items that showed good stability across both time points (i.e. had significant correlations in both “Right Now” and “Past Week” correlation matrices) were considered for analysis.

Exploratory structural equation modeling (ESEM) (77) with repeated time points (for “Right Now” and “Past Week”) was employed using Mplus version 8.4 (Los Angeles, CA) (78). The ESEM technique blends exploratory factor and confirmatory analysis strategies by flexibly capturing underlying latent constructs without *a priori* hypotheses on exactly how items will load (79). ESEM therefore has an advantage over traditional EFA as it allows for testing of specific hypotheses of latent constructs while overcoming the cross-loading constraints in CFA (80). It is also recommended over traditional factor analysis for validating exercise psychology instruments (81). Two theoretical latent factors were anticipated (“desire to move” and “desire to rest”). Model fit indices for good fit were set to at least 0.9 for TLI and CFI, and less than 0.08 for RMSEA and SRMR (82-85). In order to test for reliability of the scale, McDonald’s ω was calculated based on factor loadings (86, 87). Data were also examined for multivariate outliers using log likelihood contribution plotted against latent factor scores (88). Outliers were removed and models were re-run to determine if they affected model fit. To test hypothesis 1, given satisfactory fit of the ESEM model, scores on the CRAVE were then calculated. One sample t-tests (SPSS version 26) were used to determine if participants significantly endorsed the constructs overall.

### Results

Means (SD) for NOW items ranged from 3.18 (3.06) for “veg out” (vegetate) to 5.81 (3.16) for “be physically active”. For WEEK, they ranged from 2.78 (2.49) for “be motionless” to 7.32 (2.22) for “be physically active”. See Tables 1 and 2 for descriptive statistics. NOW and WEEK items (i.e., want to “move my body” *right now* versus *in the past week*) were correlated for move (*r* values=.30 to .60) and rest (*r* values = .41-.49).

**Table 1.**
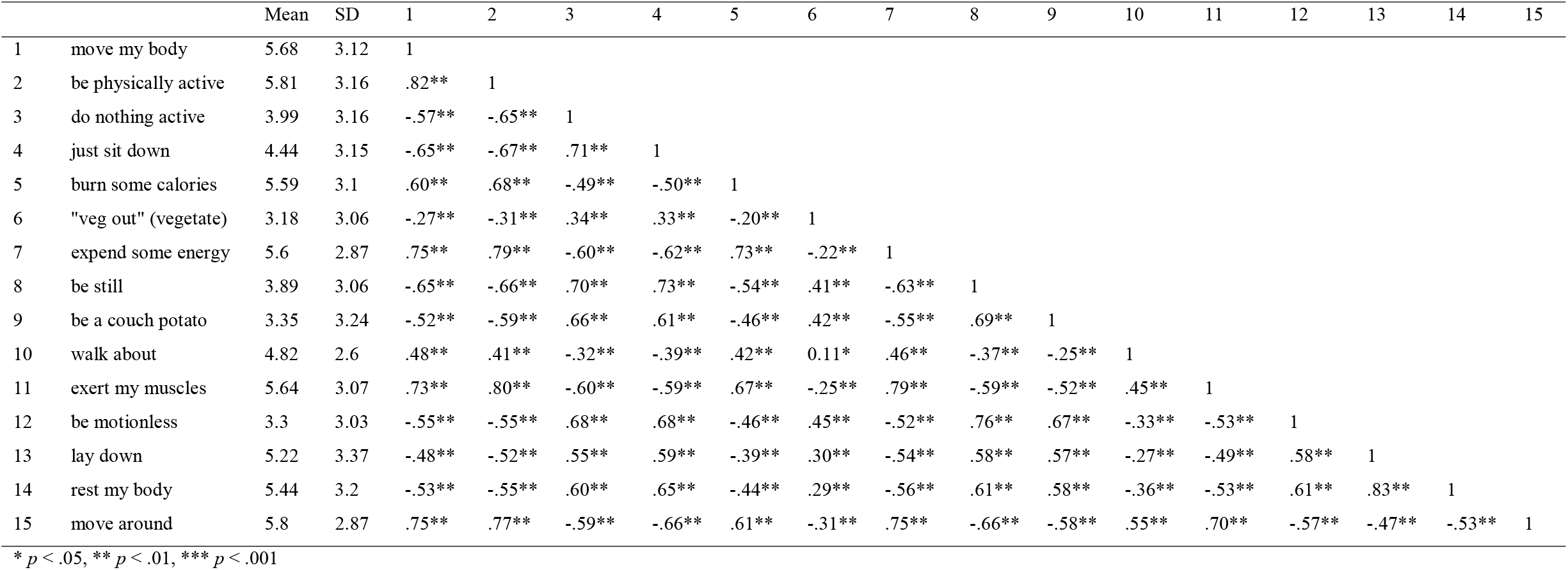
Descriptive statistics (means, SD) and inter-item correlations for CRAVE items assessed “right now” (Study 1 data)

**Table 2.**
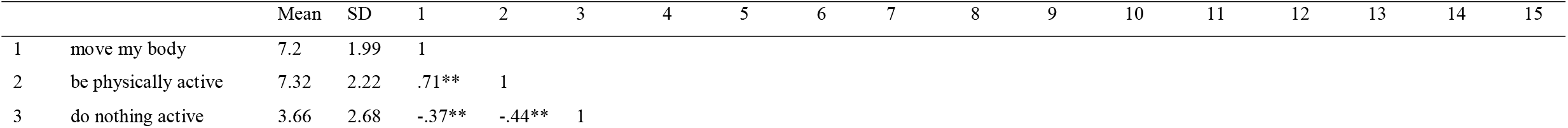
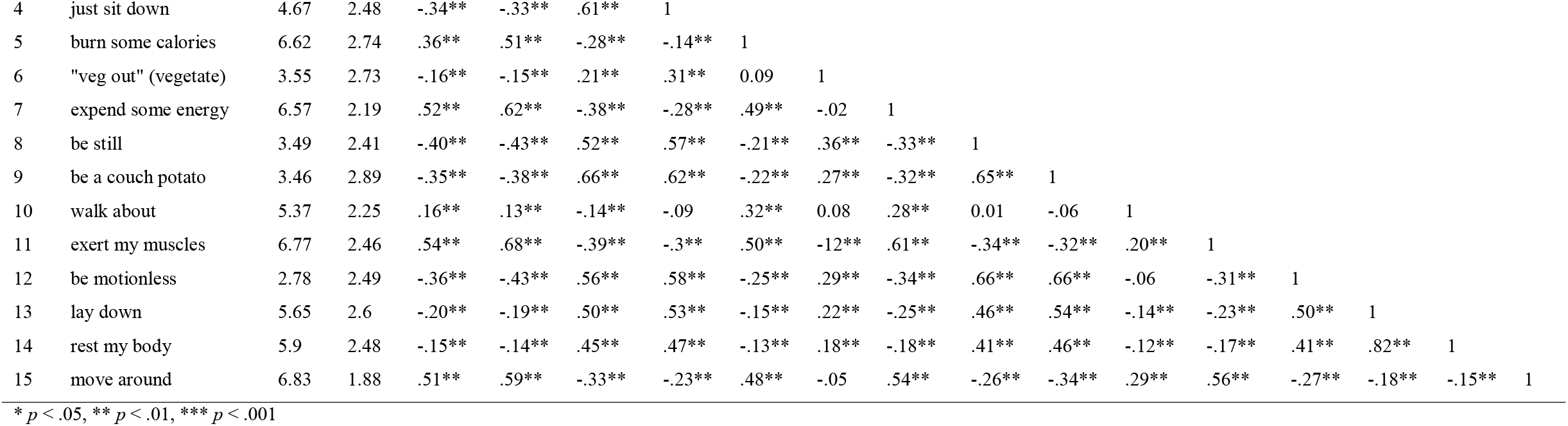
Descriptive statistics (means, SD) and inter-item correlations for CRAVE items assessed “in the past week” (Study 1 data)

Correlations amongst the “Right Now” items were all significant. However, amongst the “Past Week” version of these items, two had numerous non-significant correlations (“veg out” (vegetate) and “walk about”) and were therefore removed from analysis. Three additional items had multiple very low correlations (smaller than absolute value of 0.3) (89). These items included “burn some calories,” “lay down,” and “rest my body.” Analysis that included these items (26 in total, 13 for PW and 13 for RN) yielded a marginally poor fitting model (TLI/CFI: 0.83/0.86; RMSEA: 0.11; SRMR: 0.05). However, removal of these three items (20 in total across PW and RN) yielded a well-fitting, consistent, and parsimonious model (TLI/CFI: 0.92/0.94; RMSEA: 0.08; SRMR: 0.04). For the final model, *X* ^2^ (190) = 6516.7, *p* < .001. See Table 3 for significant loadings. Overall, “Right Now” loadings were stronger, particularly for the “Move” latent factor (.72-.92). However, both “Right Now” and “Past Week” displayed the same significant positive loadings onto the anticipated “desire to move” and “desire to rest” constructs. McDonald’s ω was very high for these items (0.97 overall), indicating good reliability of the scale. Removal of multivariate outliers slightly improved but did not substantively change model fit; thus, outliers were retained.

**Table 3A.** Study 1 Factor Loadings are standardized. Bold = significantly positively loaded *p* < 0.05; Bold italics = significantly negatively loaded *p* < 0.05

|  | Item # | Description | Factor Loading Move | Factor Loading Rest |
| --- | --- | --- | --- | --- |
| NOW | 1 | Move my body | **0.849** | -0.024 |
|  | 2 | Be physically active | **0.918** | -0.006 |
|  | 7 | Expend some energy | **0.880** | 0.005 |
|  | 11 | Exert my muscles | **0.919** | 0.066 |
|  | 15 | Move around | **0.720** | ***-0.151*** |
|  | 3 | Do nothing active | ***-0.142*** | **0.701** |
|  | 4 | Just sit down | ***-0.200*** | **0.671** |
|  | 8 | Be still | -0.104 | **0.791** |
|  | 9 | Be a couch potato | 0.004 | **0.799** |
|  | 12 | Be motionless | 0.106 | **0.930** |
| PAST WEEK | 1 | Move my body | **0.692** | -0.083 |
|  | 2 | Be physically active | **0.838** | -0.048 |
|  | 7 | Expend some energy | **0.742** | -0.003 |
|  | 11 | Exert my muscles | **0.827** | 0.049 |
|  | 15 | Move around | **0.718** | 0.029 |
|  | 3 | Do nothing active | ***-0.133*** | **0.664** |
|  | 4 | Just sit down | 0.013 | **0.744** |
|  | 8 | Be still | -0.039 | **0.766** |
|  | 9 | Be a couch potato | 0.004 | **0.828** |
|  | 12 | Be motionless | 0.003 | **0.812** |

In regards to hypothesis 1, one sample t-tests revealed perceptions of desires/wants to move and rest significantly greater than 0 (Means: Move Right Now 28.5, Rest Right Now 18.9, Move Past Week 34.7, Rest Past Week 18.0; *p*’s < 0.001), supporting the idea that people do indeed have movement and sedentary-related desires. Further supporting hypothesis 2, Move and Rest (measured “Right Now”) were correlated moderately and inversely (*r* = -.78). The “Past Week” factors for Move and Rest were more weakly correlated (*r* = -.54). In regards to hypothesis 3, the move factor assessed “right now” correlated moderately with move assessed “in the past week” (*r* = .51), and a similar correlation was found for rest (*r* = .65). Desire to move and rest scores were calculated for “Right Now” and “Past Week” based on the 10-item version (5 items per construct) by summing the raw scores on the items. When comparing Move RN vs. PW (and Rest RN vs PW), Move scores significantly differed (t(398) = 10.2, *p* < 0.001). However, Rest scores did not (t(395) = 1.4, *p* > 0.16). In other words, Move assessed right now significantly differed from move assessed in the past week, but this was not the case for Rest.

### Discussion

Study 1 initiated the development and validation of a scale to measure wants/desires for movement and rest. This process was highly structural: 30 items were generated for the scale (15 “right now”, 15 for “past week”), the items were administered to a test population of over 400 participants, the item set was trimmed, the factor structure was analyzed to determine an appropriate number of scale items to occupy each dimension, and relationships between factors were evaluated. The final scale was composed of two parts: Past Week and Right Now-each with move and rest/sedentary subscales. Each subscale, in turn, has 5 items resulting in a range of scores of 0-50 for both move and rest. Reliability of the scale was high, as determined by McDonald’s omega.

Results supported all applicable hypotheses, at least partially. Participants rated their desires to move and rest and being greater than 0 and with sufficient variability – in other words, they perceived these desires existed. This is important because some researchers have questioned the veracity of desires/wants to move as either being minimal, non-existent (90, 91) or secondary to dread/aversions for movement (15, 54). Furthermore, in line with the WANT model (17), items loaded into two distinct factors, which were moderately and inversely related. This supports the claim of the model that desires/wants for movement/physical activity are separable from desires/wants to rest or be sedentary.

The factors, originally thought of as “move” and “rest”, might be more accurately described as “move” and “sedentary”. The statistical analysis resulted in the elimination of several items originally thought to be important for the scale, including “rest my body” and “burn some calories”. Trimming these items was originally based on low inter-item correlations, but later confirmed by modelling that indicated a worse fit when these items were included. The omission of these two items is somewhat problematic as the intent of the scale was to measure aspects of “rest”. Examining the 5 remaining items (do nothing active, just sit down, be still, be a couch potato, be motionless) seems to indicate that this sub-scale is more related to sedentarism than rest, which makes sense from the standpoint that rest might be construed as a process of purposeful recovery or simply breaks from work (92). Furthermore, sleep, rest and sedentary behavior have independent effects on health and body composition (93).

Another supposition of the model is that desires/wants are transient and state-like, sometimes changing from moment to moment, but at least not being similar to traits. We asked respondents to rate their current desires and to look back and rate them over the last week. Indeed, there were differences between wants/desires for movement as reported “right now” versus in the “past week”, as determined by t tests on scale scores. However, this was not the case for rest. Evaluating desires “right now” requires being attuned to and aware of internal states (ie., interoception) (94). On the other hand, evaluating them over the past week requires retrospectively assessing fluctuations in desires over a 7 day period and ostensibly requires memory, with intensity of sensations and later sensations being in primary focus (95, 96).

There are several limitations to this study-the main ones being that it assessed a population limited to undergraduate college students who were evaluated at only 1 time point. However, these are deficiencies to be addressed in studies that follow. An important delimiter of this investigation is that we did not assess diswants or aversion/dread for movement and rest (15, 54)-choosing instead to focus on desires/wants. Despite these shortcomings, this study was the first large step in validating a scale to measure wants/desires for physical activity and sedentarism. Overall, these results indicate that the scale has good initial psychometric/structural properties and further evaluation and validity testing is appropriate.

## Study 2

### Introduction

In study 1, the factor structure of a new scale to measure desires/wants for movement and sedentarism was established, modeled and analyzed. The primary purpose of study 2 was to determine model fit in a new sample of participants, as replication of models in new samples is frequently problematic (81). A further objective was to assess the reliability of the scale and to determine if there were any issues with discriminate validity (i.e., too much overlap between move and sedentarism factors). As with study 1, these analyses allow for further examination of the WANT model (17). As such, analyses from this study are applicable to hypotheses 1-3 in the same manner as in Study 1.

Additional purposes of study 2 were to explore how the CRAVE scale varies by gender and age and determine if there is an association of move/sedentary desires with body mass index (BMI). There is a basis to believe that desires/wants to move/rest would vary by gender, as there are numerous studies indicating psychological and motivational differences between men and women (24, 97, 98). It seems plausible that those of younger age may want to move more, especially when considering movement from a lifespan perspective (97, 99-101). Those who are overweight have greater decrements in mood with small increases in workload (102), are less likely to adhere to fitness programs (103), and score higher for amotivation and externally regulated motivation (104). Thus, it seems reasonable to hypothesize that those who are more lean might have greater reinforcement for movement, and accordingly, greater desires for movement.

Desires/wants for movement may also vary by stage-of-change for exercise adoption, as specified by the Transtheoretical Model of behavior change (TTM) (105-107). The TTM posits that individuals move through five stages-of-change or readiness for health behavior, starting with precontemplation (no intention to be active) to maintenance (meeting a set level of physical activity for 6 months or longer) (105). Different sets of behavior change processes are primary in each stage, as well as the strength of various psychological attributes. There is an intersecting literature indicating that readiness, particularly a state of readiness, is related to psychological wants/desires (108). For instance, Lutz and colleagues (107) hypothesized that individuals in the maintenance stage of exercise behavior may simply be more motivated and have greater drive and impulse towards exercise. Indeed, there is some indication that desire might be more relevant for behaviors that are habitual than for those in emerging stages. In other words, desires or wants are more important for maintaining habits, while processes, such as goals, are more important for starting a new habit (109, 110). Thus, it was hypothesized that those in higher stages of change / readiness for exercise would have stronger desires/wants for movement and lower desires for sedentarism or inactivity.

### Methods

#### Participants

A total of 444 college students (*M*_age_ = 20.3 years, *SD* = 2.9) from a public Midwestern university participated. There was no overlap between studies 1 & 2 for participants. Self-report data from the demographic questionnaire highlighted the fact that 59.2% of the participants were female, 39.4% male, and 1.4% elected not to respond. The ethnic make-up of the sample was diverse; as specifically, 47.5% of the students classified themselves as Caucasian, 28.1% as African American, 8.4% as Hispanic/Latino(a), 7% as Asian American, 0.7% as Native American, 7.4% as multiple ethnicities, and 0.9% as other/not listed. Student academic rank resembled a positive skewness, with freshman accounting for 28.7% of the sample, sophomores = 29.2%, juniors = 29.8%, seniors = 11.8%, and graduate students = 0.4%. Equivalent to Study 1, 95% of participants were not involved in varsity sports at the university; however, using the stage-of-change from the Transtheoretical Model (TTM) to assess exercise engagement, 79.1% of students in the current study either exercised somewhat, exercised regularly for less than six months, or exercised regularly for more than six months. Finally, body mass index (BMI) was also calculated, revealing that the second sample had a BMI of 25.0 kg/m^2^ (*SD =* 5.2). Men were 25.9 ± 5.2, and women were 24.4 ± 5.2. All individuals signed an informed consent. The study was approved by the Institutional Review Board at Northern Illinois University, in accordance with the Declaration of Helsinki, protocol # HS13-0035.

#### Instrumentation

Similar to Study 1, participants completed a demographic questionnaire – with the only difference being: a) the inclusion of items for height and weight to calculate BMI and b) a single question to categorize Transtheoretical model (TTM) stage-of-change related to exercise involvement (107, 111). Stage-of-change was assessed using the 5-choice approach (107, 111) with each item representing the five stages. Participants were asked to select a stage based on their current exercise behavior: a) “I currently do not exercise and do not intend to exercise in the next 6 months” (pre-contemplation), b) “I currently do not exercise, but I am thinking about starting in the next 6 months” (contemplation), c) “I currently exercise some, but less than 3 times per week for 20 min of more each time” (planning), d) “I currently exercise regularly, 3 times a week or more for 20 min or more each time, *but* I have exercised this much for less than 6 months” (action), and e) “I currently exercise regularly, 3 times a week or more for 20 min or more each time, *and* I have exercised this much for more than 6 months” (maintenance). Students then responded to the identical CRAVE questions (30 items, 15 for “MOVE” and 15 for “REST”) used in Study 1.

#### Procedure

Following the procedures employed in the previous study, two researchers again contacted professors at the predetermined university who had enrollments of greater than 50 students in a class. (Note that Study 2 was conducted in the semester immediately following data collection for Study 1; thus, researchers were careful to exclude potentially large classes based on course sequencing to exclude the possibility of recurring participants.) Eight professors agreed to the request for class participation and one researcher from the byline traveled to each class for data collection. Once again, the researcher explained the nature of the study, the anonymity of responses, allowed time for questions or for students to opt out of participation (no student declined participation), distributed the questionnaires for completion before class, and collected them upon completion. Written consent was obtained from all participants.

#### Data Analysis

Congruent with Study 1, data were double entered, cleaned for extreme responders (i.e. those who answered all top or all bottom on all items), and screened for univariate outliers in SPSS version 26 (Armonk, NY, IBM Corporation). In order to confirm latent findings from Study 1, the same exploratory structural equation modeling (ESEM) (77) with repeated time points technique was employed using Mplus version 8.4 (Los Angeles, CA) (78). Both the prior identified 13-item and 10-item versions were assessed. In order to test for reliability, McDonald’s ω was also calculated based on factor loadings (86). As in Study 1, model fit indices for good fit were set to at least 0.9 for TLI and CFI, and less than 0.08 for RMSEA and SRMR (82-85). Data were also examined for multivariate outliers using log likelihood contribution plotted against latent factor scores (88). Outliers were removed and models were re-run to determine if they affected model fit.

The associations between desires/wants to move and rest and TTM stage-of-change, BMI, age, and gender were explored using multiple linear regressions in the R package (v. 3.6.1, R Core Team, Vienna, Austria). TTM stage and gender were treated as dummy coded factors while BMI and age were entered as continuous variables. Independent variables were all entered in the same step. Wants/desires to move and rest over the “past week” and “right now” encapsulated the four dependent variables. EE scores were derived by summing the 5 items identified in Studies 1 and 2 from relevant subscales. Significance was set to *p* < 0.05.

### Results

Means (SD) for NOW items ranged from 3.06 (3.06) for “veg out” (vegetate) to 6.05 (3.31) for “rest my body.” For WEEK, they ranged from 2.31 (2.39) for “be motionless” to 7.53 (2.13) for “be physically active”. Therefore, participants indicated that they did perceive desires/wants for movement and rest (hypothesis 1). See Supplemental Tables 1 and 2 for descriptive statistics and correlations for the CRAVE scale items.

Similar to Study 1 results, the 13-item version of the scale displayed a marginally poor fitting model (TFI/CFI: 0.81/0.84; RMSEA: 0.10; SRMR: 0.06). Utilizing the 10-item version previously identified in study 1 (removing “burn some calories,” “lay down,” and “rest my body”) lead to an improved, good fitting model (TFI/CFI: 0.91/0.93; RMSEA: 0.08; SRMR: 0.04). For the final model, *X* ^2^ (148) = 603.9, *p* < .001.Items loaded similarly to Study 1 (see Table 4), with interpretable “Move” and “Rest” latent variables in both the Right Now and Past Week timepoints. Removal of multivariate outliers slightly worsened model fit but not did substantively change them (all goodness-of-fit indices within desired parameters); thus, outliers were retained. McDonald’s ω was very high (0.97 overall), indicating good reliability of the scale.

**Table 3B.** Correlation matrix of latent factors. All latent factor correlations are significant at *p* < 0.001.

|  | Move<br>Now | Rest<br>Now | Move<br>Past<br>Week | Rest<br>Past<br>Week |
| --- | --- | --- | --- | --- |
| Rest<br>Now | -0.78 | -- |  |  |
| Move<br>Past<br>Week | 0.51 | -0.29 | -- |  |
| Rest<br>Past<br>Week | -0.39 | 0.65 | -0.54 | -- |

In further support of main hypothesis 2, the correlation between Rest-Move factors was inverse and moderate to strong (*r*=-.71 and *r*=-.55 for the NOW and WEEK scales, respectively), but not strong enough to suggest a significant discriminant validity issue (where *r* >.85) (112). In support of main hypothesis 3, the move factor assessed “right now” correlated moderately with move assessed “in the past week” (*r* = .52) and a similar correlation was found for rest (*r* = .65), indicating participants rated their desires/wants differently over time. Furthermore, NOW and WEEK items (i.e., want to “move my body” *right now* versus *in the past week*) were correlated for move (*r* values=.34 to .64) and rest (*r* values = .40-.49).

Multiple linear regression analyses conducted to determine the association between CRAVE constructs (10-item scale for RN and PW) and age, gender, BMI and stage-of-change for exercise were all significant: Move-RN: *F*(7, 412) = 5.763, *p* < 0.001; Rest-RN: *F*(7, 413) = 5.65, *p* < 0.001; Move-PW: *F*(7, 418) = 10.02, *p* < .001; Rest-PW: *F*(7, 416) = 2.436, *p* = 0.019. Residuals were all normally distributed. Type III tests found that exercise stage-of-change as a single variable was significant in all four models (*p* < 0.001 in all models except Rest-PW, which was p = 0.014). Therefore, stage-of-change was run as 4 categorical variables. Being the precontemplation or contemplation stages significantly predicted all four CRAVE outcomes (*p* < 0.05). Being in the planning stage was significant in 3 models (Move-RN/PW and Rest-RN; *p* < 0.05). Age only predicted Rest “Right now”, B = −0.555, SE = 0.230, *t*(413) = −2.417, *p* = 0.016. Gender and BMI did not predict any CRAVE outcome. See Table 5. the psychometric properties of the CRAVE, 400+ new participants were administered the CRAVE scale (both Past Week and Right Now). Most importantly, the final exploratory structural equation model (ESEM) utilized in Study 1 was replicated with a good fit, indicating the model has good stability across two independent samples (83). Furthermore, reliability (internal consistency) of the subscales was very good. Factor loadings generated from the ESEM supported the current model structure and move/rest factors were moderately and inversely related. Thus, problems with discriminate validity were not detected. “Right now” and “Past week” factors were moderately correlated for both move and rest, indicating perceived changes in wants/desires over time. Consequently, it appears that objectives in regards to the scale as well as hypotheses specific to the WANT model were all supported.

**Table 5.**
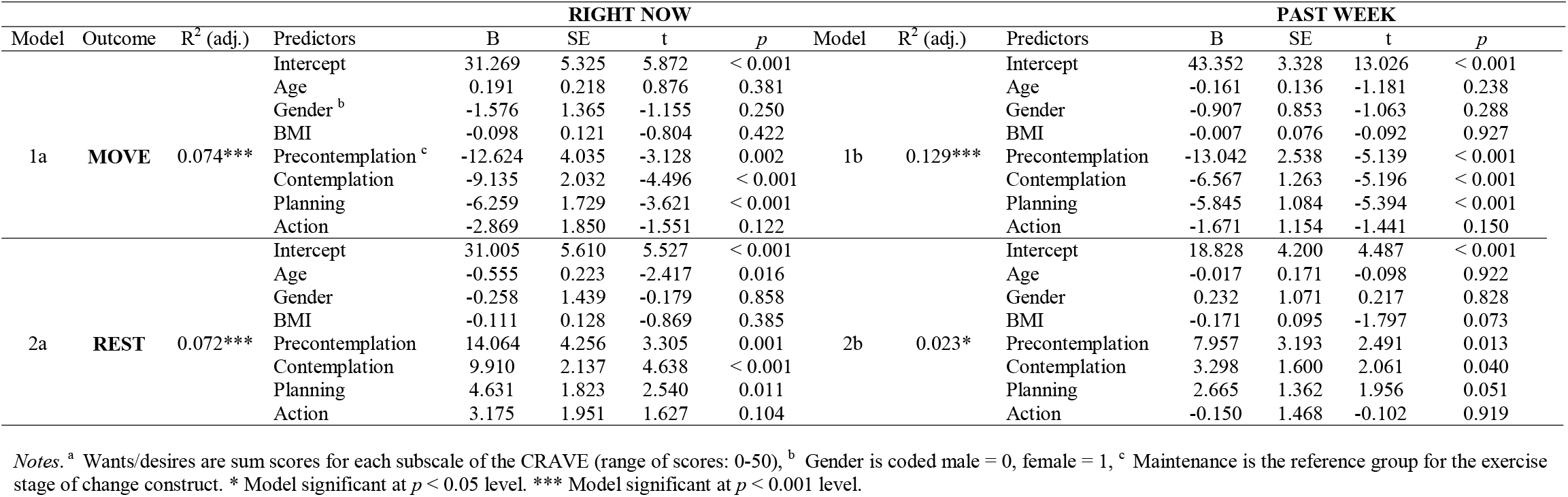
Associations of age, gender, BMI and exercise stages of change (Transtheoretical Model) with desires/wants to move and rest, both “right now” and “in the past week”^a^

The most consistent finding from the multiple linear regression analyses was the association of exercise stage-of-change with desires/wants to move and rest. Compared to those in the maintenance stage (consistently exercising for 6 months), those who classified themselves as pre-contemplators (not considering exercise), contemplators (those thinking about starting exercise) and planning to start an exercise regimen rated their desires to move as lower and their desires to rest as higher. The same trend was observed for those in the action stage (exercising inconsistently), though the relationship was not significant. Thus, one might infer that those most habituated to exercise have the strongest desires to move and the lowest desires to be sedentary. This seems to be consistent with observations that wants/desires might be more important for behavior in those who are habitual exercisers (107, 109, 110). On the other hand, cognitive approaches (e.g., goals, intentions) may be more important targets for those wanting to initiate an exercise regimen. It is also possible those in lower stages of change have greater competing desires (e.g., from other pleasurable stimuli), notice their desires for movement less (i.e.., are less attentive) are more sensitive to desires to be sedentary (17) and/or are dampened by external forces or internal states, such as the experience of stress (107). Lesser desires to move and greater desires to rest might contribute to poorer regulation of physical activity in lower stages of change (113), and these data further support the notion that a stage-based approach to behavioral counseling is useful (114). Such questions deserve greater attention.

Against expectations, gender, age and BMI were largely unrelated to CRAVE scores. Age was linearly related to rest “right now”, with those who were older having less of this desire. However, age was unrelated to other aspects of desire/wants for movement and rest. Perhaps those who have been in college longer have better sleep regulation (i.e., less sleep debt) or are less disrupted by the transition from home to college (115). Or perhaps they are less impacted, noticing their desires less or managing them better (116). It is also possible that younger college students were taking earlier classes (e.g., 8am) due to university course registration policies. Unfortunately, we could not conduct an analysis by time of day. It would be useful to investigate differences in CRAVE scores among college class levels (i.e., freshmen, sophomores, graduate students).

This study had several limitations that should be considered in future studies. First, the sample included in this study (and in study 1) had a limited range for age and BMI. Inquiry into exercise behavior was limited to stage-of-change, and other aspects of physical activity and exercise behavior should be considered (4). We also did not measure any aspect of sleep or resting behaviors, like napping, which are ostensibly impactful on desires to rest and be sedentary and vice versa (117). This investigation did not control for time-of-day (e.g., morning, afternoon) or seasonal effects, which likely relate to exercise behavior and motivation to move (118). The issue of time of day is considered in the following studies below, however. Inclusion of such variables might account for a much greater proportion of the variance in CRAVE scores, whereas in the current models, only a small amount of variance was explained. Despite these shortcomings, results from Study 2 provide further evidence that the CRAVE has good psychometric properties and may be moved forward for further validity testing.

## Study 3

### Introduction

The main purpose of Study 3 was to investigate the reliability of the Move and Rest subscales of the CRAVE tool throughout a 2-year time span, as well as within the same day (2 points, pre and post of a laboratory session). Study 3 directly tested hypothesis 3 and supported hypotheses 1 and 2. Specifically, we hypothesized that Move and Rest scores would show greater reliability within the same day than across 2 years. These findings would suggest that the desire to move or rest has state-like properties rather than trait-like properties. States are brief, temporary, and easily influenced by changes in the external environment or internal milieu. Meanwhile, traits are stable, long-lasting, and emanate primarily from within the person (119). A secondary aim, not in the main hypotheses, was to determine which desire was rated stronger -move or desire. This has never been tested before, but one might surmise that humans desire or want rest (including sleep and relaxation) more than movement (including leisure time physical activity and exercise). Hoffmann (12, 43) found that desires to sleep/rest are the most common desires humans have and outrank other desires, such as sports participation.

### Methods

The CRAVE Scale was administered to 127 participants (28.1±7.9 yrs, 47% female) at 0, 6, 12, 18 and 24 months. The 13-item version for “Right now” was utilized as found from Studies 1 and 2 (10 items scored and 3 are fillers). The ethnic make-up of the sample was diverse, as 42.5% of participants classified themselves as Caucasian, 32.3% as African American, 8.7% as Hispanic, 7.9% as Asian, 7.9% as Other and 0.8% did not report ethnicity. Participants were queried at two time points (i.e., Point 1; Point 2) at baseline and each of the four follow-up sessions. CRAVE scores were collected before and after they completed a battery of self-report and structured interviews that were part of a larger study aimed at understanding the motivation to eat hyperpalatable foods (120).

Statistical tests were performed using RStudio (Version 1.2.1335; R Foundation for Statistical Computing, Vienna, AT). Participants completed the CRAVE a variable number of times. Therefore, within subjects test-retest reliability was assessed with an inter-class correlation (ICC) generated with a random intercept model using a linear mixed effects model. Test-retest reliability was assessed for within session (i.e. Points 1 and 2 of the testing session) and between sessions (i.e. at timepoints 0, 6, 12, 18, and 24 months) for Move and Rest subscores. To ascertain the effects of time, both across sessions (Points 1 and 2) and across months (0-24), a linear mixed effects model was created with 3 terms: session time point, month and an interaction term. All individuals signed an informed consent. The study was approved by the Human Investigation Committee at the Yale School of Medicine, in accordance with the Declaration of Helsinki.

### Results

On average, participants completed the CRAVE 5.4 ± 3.04 times over 24 months. Because the scale was introduced as part of an ongoing clinical trial, the number of observations (Point 1 / Point 2) at baseline and 6, 12, 18 and 24 months were 50/36, 61/59, 68/67, 78/74 and 97/96, respectively. Descriptive statistics and within-day correlations and intra-class correlations are presented in Table 6. Move was significantly higher than rest for time 1 of the lab session (25.08 ± 10.9 vs. 17.6 ± 10.8, *p* < .001). Move was also higher than rest for time 1 (end of session) (24.52 ± 11.3 vs. 16.45 ± 11.0, *p* < .001). In fact, Move scores were consistently higher than Rest scores at each time point across all 24 months. See Figure 1. Move scores taken within the same day (Points 1 and 2) had ICC’s = .72-.95, and rest scores taken within the same day had ICC’s = .69-.89, indicating at least moderate strength (121). However, across 24-months, move (ICC = .53) and rest scores (ICC = .49) had lower reliability. Therefore, the CRAVE Scale showed greater test-retest consistency within each day than across 0, 6, 12, 18 and 24-months. A linear mixed effects model revealed that there was no significant effect of session time point (pre/post), but month did trend for significance for both move and rest (Move: (*F*(309.69, 1) = 3.6425, *p* = .057); Rest: (*F*(314.10, 1) = 3.4905, *p* = .063) The interaction term between CRAVE Scores measured across months and within each day was not significant Move: (*F*(318.11, 1) = 0.4113, *p* = .52); Rest: (*F*(328.55, 1) = 0.0004, *p* = .98).

**Table 6.** Means, standard deviations, Pearson’s correlations (r), and intra-class correlations ^a^ (ICC) for Move and Rest Scores within the same test day (session points 1 and 2) across two years of follow-up ^b^.

| Time (months) | Move (Point 1) | Move (Point 2) | Move Scores (r) | Move Scores (ICC) | Rest (Point 1) | Rest (Point 2) | Rest Scores (r) | Rest Scores (ICC) |
| --- | --- | --- | --- | --- | --- | --- | --- | --- |
| 0 | 27.4 ± 12.7 | 29.9 ± 10.9 | 0.93 | 0.95 | 17.0 ± 10.1 | 13.8 ± 8.7 | 0.83 | 0.89 |
| 6 | 23.8 ± 10.6 | 22.4 ± 11.9 | 0.74 | 0.72 | 16.2 ± 10.3 | 16.4 ± 11.8 | 0.77 | 0.69 |
| 12 | 25.5 ± 11.5 | 24.2 ± 11.4 | 0.80 | 0.79 | 16.8 ± 10.3 | 16.3 ± 11.2 | 0.88 | 0.86 |
| 18 | 24.2 ± 11.0 | 24.9 ± 11.3 | 0.86 | 0.85 | 19.0 ± 11.2 | 16.7 ± 10.7 | 0.75 | 0.75 |
| 24 | 25.1 ± 9.6 | 23.7 ± 9.8 | 0.82 | 0.82 | 18.2 ± 11.3 | 17.4 ± 11.4 | 0.84 | 0.86 |

<sup>a</sup> ICC's are derived from a linear mixed effects model that has a random intercept for each participant.
<sup>b</sup> Move trended higher over two years ( $p = .057$ ) and rest trended down ( $p = .063$ ).

**Table 6A.** Study 2. Factor Loadings are standardized. Bold = significantly positively loaded *p* < 0.05; Bold italics = significantly negatively loaded *p* < 0.05

|  | Item # | Description | Factor Loading Move | Factor Loading Rest |
| --- | --- | --- | --- | --- |
| NOW | 1 | Move my body | **0.849** | -0.046 |
|  | 2 | Be physically active | **0.918** | -0.007 |
|  | 7 | Expend some energy | **0.880** | 0.045 |
|  | 11 | Exert my muscles | **0.919** | 0.062 |
|  | 15 | Move around | **0.720** | -0.025 |
|  | 3 | Do nothing active | **-0.142** | **0.653** |
|  | 4 | Just sit down | **-0.200** | **0.667** |
|  | 8 | Be still | -0.104 | **0.886** |
|  | 9 | Be a couch potato | 0.004 | **0.816** |
|  | 12 | Be motionless | 0.106 | **0.875** |
| PAST WEEK | 1 | Move my body | **0.737** | -0.012 |
|  | 2 | Be physically active | **0.832** | -0.002 |
|  | 7 | Expend some energy | **0.734** | 0.004 |
|  | 11 | Exert my muscles | **0.772** | 0.060 |
|  | 15 | Move around | **0.674** | -0.043 |
|  | 3 | Do nothing active | -0.093 | **0.680** |
|  | 4 | Just sit down | 0.024 | **0.717** |
|  | 8 | Be still | **0.146** | **0.819** |
|  | 9 | Be a couch potato | -0.031 | **0.711** |
|  | 12 | Be motionless | -0.008 | **0.788** |

**Table 6B.** Correlation matrix of latent factors. All latent factor correlations are significant at *p* < 0.001.

|  | Move<br>Now | Rest<br>Now | Move<br>Past<br>Week | Rest<br>Past<br>Week |
| --- | --- | --- | --- | --- |
| Rest<br>Now | -0.713 | -- |  |  |
| Move<br>Past<br>Week | 0.518 | -0.302 | -- |  |
| Rest<br>Past<br>Week | -0.335 | 0.654 | -0.554 | -- |

**Figure 1.**
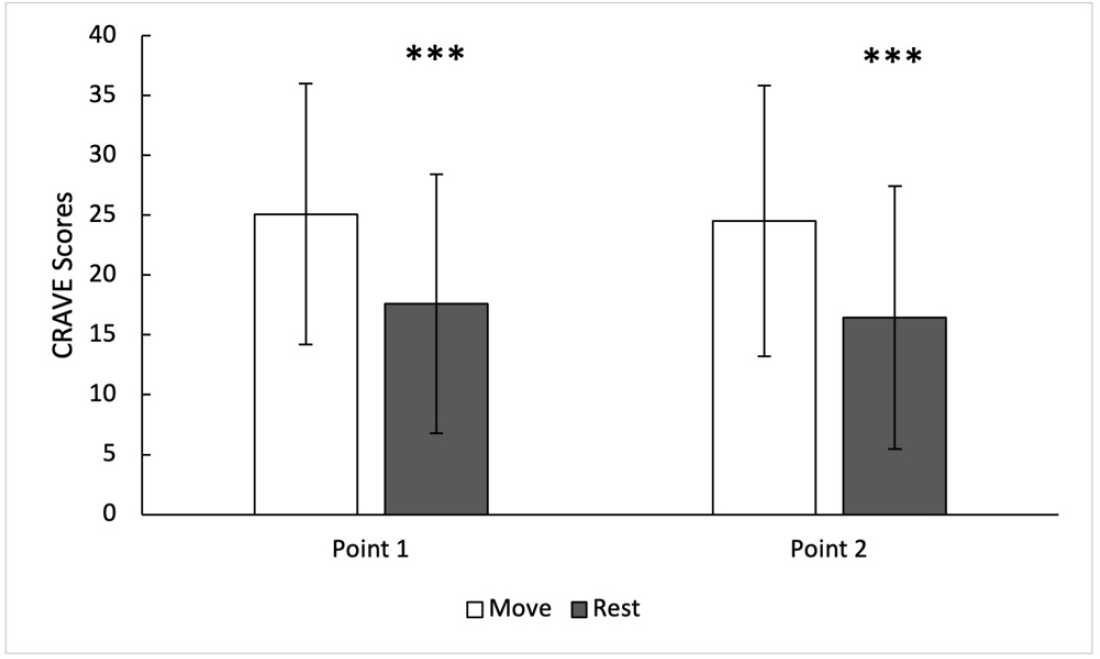
Desires/wants to move and rest at Point 1 (pre-lab session) and Point 2 (post-lab) across 24 months of follow up. *** denotes *p* < .0001

### Discussion

The results from Study 3 support the third general hypothesis, that desires to move and rest are transitory and have state-like qualities. This was demonstrated by associations that were significantly greater between the urge to move and rest at two points within the same day compared to associations between these desires across multiple months. In other words, there was greater reliability within the same day than across 2 years. These findings suggest that the desire to move and rest have state-like properties rather than trait-like properties and are congruent with previous literature demonstrating that the desire to be physically active is not stable and can be affected by a myriad of fluctuating psychological factors, such as mood, affect, state anxiety, anger, and stress (17, 122). Furthermore, Rowland has proposed that the desire to perform physical activity can be affected by proximal factors such as personal desires, peer influences, and environmental conditions (45) as well as dietary and pharmacological influences (17).

Interestingly, desires to move and rest did not vary across the laboratory session (Points 1 and 2), but there was a trend for move to increase and rest to decrease across two years of time. Such trends might be due to factors, such as aging (97) and key experiences (101, 115). It could also be caused by reactance and acclimation to the scale itself. Some researchers remove the baseline responses from analyses for this reason (107). [Note: Evidence presented later in Study 5 seems to negate this possibility.]

Of note, participants rated their desires to move higher than their desires to rest at every query point in the study. No formal hypothesis was formulated in regards to this finding, but the trend was readily apparent. From an evolutionary standpoint, this might make sense, as movement is required for daily function in life, such as acquiring food, seeking shelter, and play, and behaviors that have high utility are likely to be wanted (17, 123). On the other hand, behavior, such as structured exercise, has only recently been reinforced in human history (124). Likewise, Hoffmann (12, 43) has found that desires for rest/sleep are the most common desires that participants have, outweighing desires, such as food, coffee, sex, and sports participation. One might consider that the finding of higher desires for move compared to rest is due to sampling bias. However, a strength of this study is that it included >600 observations of the CRAVE scale, measured over 2 years in 125 participants from a community-residing sample.

## Study 4

### Introduction

The purpose of Study 4 was to investigate changes in the desire to move and rest after a bout of maximal exercise. Indeed, a hallmark of any urge or craving is change in intensity across time, which sometimes occurs rapidly (43, 125). In particular, there is typically a reduction in desire once a sufficient quantity or intensity of a relevant stimulus is experienced, in other words, satiation is reached (125). Conversely, during deprivation, desires will increase. Concomitantly, at sufficiently high levels of either satiation or deprivation, the experience may be discomfort and aversive sensations (e.g, feeling “full” or “stuffed” after overeating) (125, 126). It seems reasonable that the same should hold true for desires for movement and rest in response to varying levels of physical activity and sedentary behavior.

A maximal treadmill test provides an opportunity for both an excess of movement and the physiological deprivation of rest. Study 4 directly tested hypotheses 3, 4, and 5 (that movement and rest desires are states, change with stimuli and are related to psychosomatic sensations). We hypothesized that a bout of maximal exercise will satiate the desire to move and deprive the body of respite, thereby leading to reductions in the desire to move and increases in the desire to rest. Maximal exercise results in considerable increases in pain and fatigue and large decreases in perceived energy (127-129). Such unpleasant sensations seem likely to result in reductions in desire to move, increases in desire to rest and/or increases in aversions or dread to move (17). We hypothesized, in accordance with hypothesis 5, that desires/wants to move and rest will be related to, but distinct from sensations of physical and mental energy and fatigue. This will also help to establish the convergent and discriminate validity of the CRAVE scale (68, 69).

### Methods

Study 4 included 21 students (*M*_age_ = 20.5±1.4 yrs; 58% female) who identified themselves as 42.9% Caucasian, 19% Asian/Pacific Islander, 19% Hispanic, 14.3% multiple races and 4.8% Arab. The students were mainly undergraduates participating in physical activity classes at The University of Texas at Austin (Austin, TX, USA). The study was approved by the University of Texas at Austin’s Institutional Review Board and informed consent was obtained from all participants.

Participants were advised to refrain from exercise for 48 hours prior to the laboratory testing session. During the testing session, participants completed a maximal, graded treadmill test that was developed for this population of college students. In this protocol, the grade or speed of the exercise was increased every minute for the first 10 minutes and then speed only was increased every minute until participants reached volitional fatigue. Once this demarcation was reached, speed and grade were reduced to provide the participant with a 5-minute cool down walk.

The CRAVE, physical energy, physical fatigue, mental energy and mental fatigue were measured one minute prior to starting the treadmill test, as well as two minutes after completing the test. The CRAVE 13-item version for “Right now” was utilized as found from Studies 1 and 2 (10 items scored and 3 are fillers). Physical and mental fatigue and energy were measured pre- and post-exercise using Visual Analogue Scales developed and validated by O’Connor (75, 130). Respondents placed a mark on a standard 10-cm line. Examples of anchors included, “I have no energy” to “Strongest feelings of energy ever felt”. Sense of effort was assessed with the Rating of Perceived Exertion (RPE) scale from Borg (131) with a range of 6-20. Statistical tests were conducted using SPSS (Version 25; IBM SPSS Statistics Software, Chicago, IL, USA). Paired t-tests were used to compare pre- and post-CRAVE scores. Effect sizes were calculated using Cohen’s d_av_ (132, 133), as well as conditional R^2^, which is intended to obtain an effect size that considers fixed and random effects in a mixed-effects model (134). Pearson’s correlations were used to assess relationships between all CRAVE scores, mental fatigue and energy, as well as physical fatigue and energy.

### Results

On average, participants took 13.5±2.0 min to reach volitional fatigue on the treadmill test, reached a maximum heart rate of 194.7 ± 10 bpm and reported a RPE of 18.3±1.0. Desire to move significantly decreased from pre-to post-treadmill test (33.5±8.3 vs. 24.8±8.3, *p <*.01, Cohen’s d_av_ = 1.05, R^2^ = .62), as shown in Figure 2. Conversely, desire to rest significantly increased from pre-to post-treadmill test (11.7±12.8 vs. 20.4±8.5, *p <* .01, Cohen’s d_av_ = .82, R^2^ = .30). Baseline desire to rest was significantly associated with the change in move (*r = 0*.*41, p* < .01) and change in rest (*r =* -.43, *p <* .01). In addition, change in desire to move was inversely associated with change in desire to rest (*r =* -.69, *p <* .001). Pre-exercise move and rest were inversely associated (*r* = -.37, *p* = .01), and the inverse relationship between move and rest scores was stronger post-exercise (*r* = -.64, *p* <.001).

**Figure 2.**
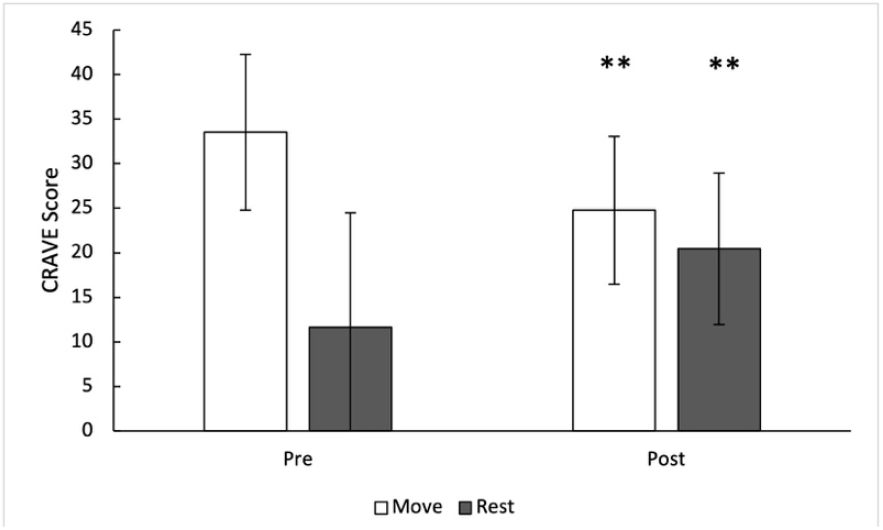
Desires/wants to move and rest pre- and post-maximal treadmill exercise test Rest is a 5-item scale with a score range of 0-50; Move is a 5-item scale with a score range of 0-50 denotes *p* < .05; ** denotes *p* < .01

Figure 3 shows the results of all correlation tests conducted. Change in desire to move had a negative association with change in physical fatigue (*r =* -.50, *p* < .001) and with change in physical energy (*r =* .33, *p* < .01), but not with mental energy (*r =* -.09) or mental fatigue (*r =* -.09). Change in rest had a significant inverse correlation with change in physical energy (*r =* -.62, *p <* .001) and a positive correlation with change in physical fatigue (*r* = .51, *p* < .001). However, it was not correlated with change in either mental energy or fatigue.

**Figure 3.**
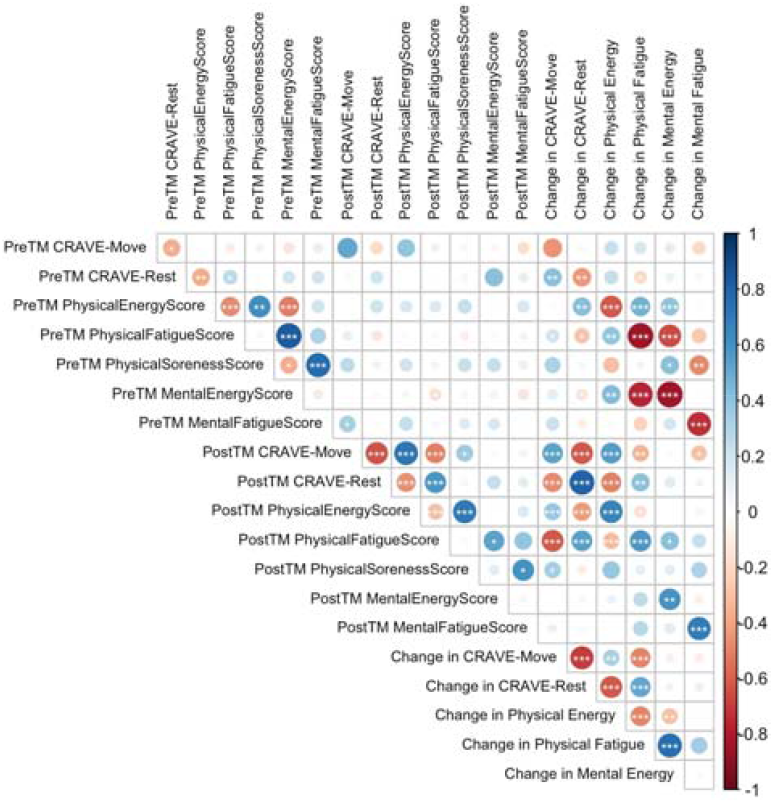
Pearson correlation matrix between pre- and post-treadmill test move and rest scores, physical energy, physical fatigue, mental energy and mental fatigue scores, and change in those scores. TM = Treadmill * denotes *p* < .05; ** denotes *p* < .01; *** denotes *p* < .001

### Discussion

Data from Study 4 demonstrated that desires to move and rest change with a maximal exercise stimulus, thus supporting hypotheses 3 and 4. Desires/wants to move and rest appear to function as psychological states that are sensitive to the provision of an exercise stimulus. In accordance with the WANT model (Stults-Kolehmainen, et al., 2020), a bout of maximal exercise appears to satiate the desire to move and places the body in a state of physiological deprivation for rest. This was demonstrated with a decrease in desire to move and an increase in desire to rest from pre- to post-treadmill exercise. Further, these findings show that perceived physical fatigue has a positive association with the desire to rest, and an inverse association with the desire to move. Additionally, perceived physical energy had a negative association with the desire to rest. Correlations were small to medium in size, providing evidence that the CRAVE captures a unique construct. Overall, these findings provide convergent validity of the scale. They also support the use of the CRAVE Scale as a tool that is sensitive to changes in the desire to move and rest before and after exercise.

Several strengths and limitations were identified. A limitation to this study is that the pre-test time period was not monitored for activity, and it is not known whether participants honored the request to refrain from exercise for 48 hours. Therefore, it is unknown if the desire to move was sufficiently built up during this time. Also, the current study did not assess positive and negative affect – other factors that likely relate to desires to move, but not strongly (20). A strength of this investigation was that all participants were pushed to their apparent physiological maximum during treadmill exercise. Everyone followed a controlled protocol with no premature conclusions. However, this was not verified with an analysis of inspired and expired gases (135). In a future study, researchers should examine changes in the desire to move and rest when completing an exercise protocol where everyone achieves a set relative intensity for a given time period (e.g., 15 minutes at 85% of VO_2max_). It would also be useful to examine these changes with a more ecologically valid exercise protocol, in a real-life exercise setting such as a full resistance training workout, sports game, a 2-mile run, etc. Future studies should also track recovery of desires for a sufficient period after they have been altered (e.g., over 30 minutes, 2 hours or more) (128, 136).

## Study 5

### Introduction

In Study 4, we investigated changes in the desire to move or rest in response to a bout of maximal exercise. In contrast, the purpose of Study 5 was to assess changes in the desires to move or rest in response to sedentary behavior. Study 5 directly tested hypotheses 3 and 4 (the desires will change in a state-like manner with the avoidance of movement), hypothesis 5 (relation to psychosomatic sensations), and indirectly tested hypotheses 1 and 2 (desires for movement exist and are separate from desires to rest). Desires to move and rest should vary by deprivation or satiation of these behaviors (125). Consequently, restricting movement, particularly in a group of individuals more likely to move (i.e., young adults) should result in increased desires to move. Conversely, satiating the need to rest through sedentary behavior should result in fewer of these desires. As the sedentary behavior is prolonged, the desire to move will build up and be felt as a type of tension, perhaps analogous to appetite (9, 45, 137). Based on these premises, we hypothesized that prolonged sitting during a university lecture period would increase the desire to move and decrease the desire to rest (hypotheses 3 and 4). Convergent and discriminate validation of the CRAVE scale requires examining other instruments that are also psychological states that may overlap but be distinct from desires to move and rest (68, 69). It is proposed that desire to move/rest will be related to, but distinct from, perceptions of energy, fatigue, tension and calmness (in accordance with hypothesis 5). A final aim was to determine the test-retest reliability of the CRAVE scale.

### Methods

The CRAVE Scale and Thayer Activation-Deactivation (AD) Checklist (138) were administered to 41 students (mean age 22.5±5.1 years; 24.4% female) before, during and at the end of a 50-minute lecture. The 13-item version for “Right now” was utilized as found from Studies 1 and 2 (10 items scored and 3 are fillers). In this study, 73.2% of participants identified themselves as Caucasian, 12.2% as Multiple Races, 2.4% as Native American, 2.4% as Hispanic, 2.4% as African American, 2.4% as other and 4.9% did not report ethnicity. Students quietly sat, listened, and took notes while the instructor delivered a PowerPoint presentation. The AD Checklist consists of 20 items answered on a 1-10 Likert Scale and measures perceived energy, tiredness, tension and calmness. This measure was only assessed pre-lecture. Lectures were at either 9AM, 11AM or 3PM, when one of the current investigators (AD) was scheduled to provide classroom instruction. Statistical tests were conducted using RStudio (Version 1.2.1335; R Foundation for Statistical Computing, Vienna, AT) and SPSS (Version 25; IBM SPSS Statistics Software, Chicago, IL, USA). A linear mixed effects model was used to compare pre-, mid- and post-lecture CRAVE Scores. Correlations were calculated to evaluate CRAVE and AD Checklist relationships. One-way ANOVA tests were used to assess differences between class times for Move and Rest Scores at the pre-, mid-, and post-lecture conditions. Reliability index was assessed with inter-class correlations, where the reliability of the pre-, mid-, and post-lecture move and rest scores was considered. All individuals signed an informed consent. The study was approved by the Institutional Review Board at Western Illinois University, in accordance with the Declaration of Helsinki.

### Results

It was found that the move scores were significantly higher post-lecture compared to pre-lecture (26.8±10.8 vs. 22.4±11.3, *p* < .01) and trended towards a significant difference compared to mid-lecture (23.5±11.3, *p* = .051), as shown in Figure 4. A main effect of time was found for rest scores (F(2,78) = 3.76), *p* < .05), where post-lecture rest scores were significantly different from mid-lecture (18.0±11.9 vs. 21.8±13.4, *p* < .05), but not significantly different from pre-lecture scores (21.0±12.2, *p* = .11). Inter-class correlations were of moderate strength for the desire to move (*ICC* = .68) and the desire to rest (*ICC* = .72) (121).

**Figure 4.**
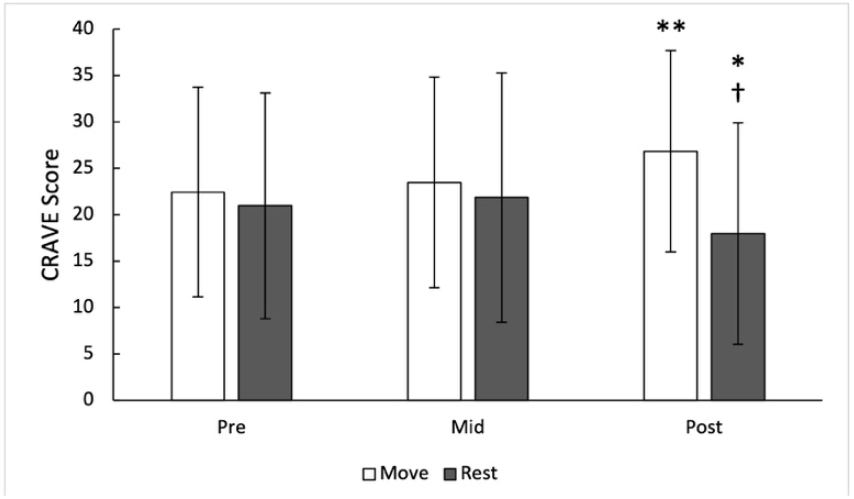
Desire to move and rest pre-, mid- and post-lecture Rest is a 7-item scale with a score range of 0-70; Move is a 6-item scale with a score range of 0-60 * denotes *p* < .05; ** denotes *p* < .01; † denotes significant difference from mid- to post-lecture

There were no differences in Move or Rest scores based on time of day for any of the pre-, mid-, or post-lecture conditions. As seen in Figure 5, pre-lecture move scores were positively associated with mid-lecture move scores (*r* = .66, *p* <.001), post-lecture move scores (*r* = .56, *p* < .01), pre-post changes in rest scores (*r* = .59, *p* < .01) and inversely associated with pre-lecture rest scores (*r* = -.62, *p* <.001), mid-lecture rest scores (*r* = -.52, *p* < .01), post-lecture rest scores (*r* = -.18, *p* < .05). Pre-lecture rest scores were positively associated with mid-lecture rest scores (*r* = .68, *p* <.001), post-lecture rest scores (*r* = .70, *p* < .001), and inversely associated with both mid-lecture move scores (*r* = .52, *p* < .001) and post-lecture move scores (*r* = -.58, *p* < .001). Pre-to-post changes in move scores were negatively associated with pre-to-post changes in rest scores (*r* = -.68, *p* <.001). Pre-lecture move scores had small and inverse associations with pre-lecture tiredness (*r* = -.17, *p* < .05) and total deactivation (*r* = -.20, *p* < .05). Mid-lecture move scores were also inversely associated with pre-lecture tiredness (*r* = -.31, *p* < .01) and total deactivation (*r* = -.35, *p* < .05).

**Figure 5.**
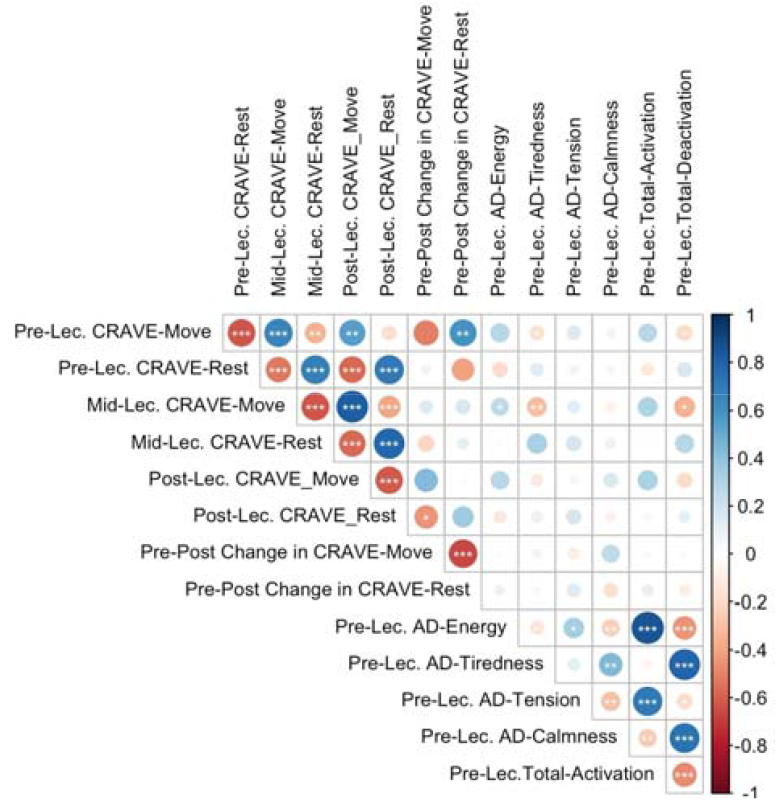
Pearson correlation matrix between pre, mid-, and post-lecture move and rest scores, energy, tiredness, tension, calmness, total activation and total deactivation. Lec = lecture; AD = Thayer Activation-Deactivation Checklist. * denotes *p* < .05; ** denotes *p* < .01; *** denotes *p* < .001

When the CRAVE was scored with all 13 items (7 for rest, 6 for move), in other words, scoring with the inclusion of the filler items, there were some additional significant findings. There was a significant difference between pre- and post-lecture for Rest (33.4±2.8 vs. 28.3±2.8, *p* < .05). Further, there was a significant difference between mid- and post-lecture move scores (28.5±2.0 vs. 32.2±2.0, *p* < .05). There were also four additional significant correlations with the Activation/Deactivation Checklist: pre-lecture move score was significantly associated with energy (*r* = .38, *p* < .05) and calmness (*r* = -.47, *p* < .01). Pre-lecture rest was significantly associated with energy (*r*= -.38, *p* < .05) and tiredness (*r* = .48, *p* < .01).

### Discussion

It was found that the desires to move and rest change throughout the course of a period of sedentary behavior. Specifically, prolonged sitting during a university lecture was associated with an increased desire to move and decreased the desire to rest. This seems to align with literatures that describe movement restriction and prolonged sitting in the classroom and in airplanes as resulting in greater somatic complaints (139), discomfort (140), and inability to attend to instruction (141). Alternatively, forthcoming movement transitions and the anticipation of movement may generate desires to move and allay those for rest. Desires to rest and move, as measured by the 10-item version of the CRAVE, were related to tiredness but not other psychosomatic sensations. Likewise, Casper has found that urges to move, as retrospectively assessed over two weeks, were unrelated to physical and mental tiredness and energy (52, 60). Data from Study 5 support the notion that desires are states. Accordingly, the CRAVE demonstrated “moderate” reliability, as might be expected from the beginning of a class to the mid-point and the end. Overall, these findings show that the CRAVE Scale is a tool that is sensitive to changes in the desire to move and rest before and after a relatively short period of sedentary behavior.

This study had a number of strengths and limitations. A strength of this investigation was that the observation periods were standardized across the sedentary period (pre-, mid- and post-lecture), and the data was collected at multiple times during the day. However, students did not return to be observed at multiple hours of the day; therefore, within-participant difference at each time of day could not be assessed. Moreover, no data was collected in the evening or at awakening. Consequently, this methodology is limited in the extent to which it can ascertain the influence time of day has on changes in the desire to move or rest. Future research should track individuals multiple times per day, on weekdays and weekends, from the moment of waking until bedroom to understand the nature variation of desires to move and rest throughout a day and across a week or longer. It would be useful to collect these with sensor data and construct models to understand covariates of the effects, such as physical activity, diet, sleep and the environment (142).

### General Discussion

The main objective of the current investigation was to create and validate an instrument to measure affectively-charged motivation states (ACMS; e.g., desires, wants, urges) for movement and rest, which was named the CRAVE. Exploratory structural equation models run in two samples revealed that a two-factor solution for movement (5 items) and rest (5 items) desires/wants produced the best fit. These two factors were inversely correlated at a moderate level, suggesting an overlap but no issues with discriminant validity. Results across studies indicated that desires/wants are transient and typify a motivation state rather than a trait. The CRAVE was sensitive to change when participants partook in a maximal treadmill test as wants/desires for rest greatly increased and desires for movement decreased. On the other hand, changes were much smaller across an hour-long period of sitting and listening. Responses to the scale were more closely correlated when observations were separated by just a few hours, compared to intervals of 6 months or more. Interestingly, desires to move were consistently higher than desires to rest. Overall, these studies found substantial support for the psychometric properties of the CRAVE, including reliability, and they also supported tenets of the WANT model.

The current data indicate that desires for movement and rest likely fall on two different, but related factors. In other words, the desire for movement is not simply *the lack of desire* for rest, or sedentary behavior, which would imply that the two desires are opposite poles on the same dimension (17). A two-factor distinction mirrors current thinking about physical activity and sedentarism, as introduced above (2, 3). Furthermore, it may be possible that one has high desire for both rest and movement simultaneously, or low desire for both as well. Being high on desire for movement does not necessitate a low level of desire for rest; as in fact, the two may be in conflict. Unfortunately, it was not within the scope of the current studies to specifically test for various patterns of responses, but a more expansive review was conducted by Stults-Kolehmainen (17). From a neurological perspective, regulation of impulses in the brain for energy expenditure and sedentary behavior appears to fall on two distinct corticostriatal pathways, which Beeler and colleagues call “Go” and “No Go” (8). Furthermore, brain systems responsible for sensations of reward and sensations of pain regulate both aversion to movement and/or “craving” for physical activity, and these differ from systems responsible for desire for rest (6). These models are also parsimonious with a vast literature base describing function of the sympathetic and parasympathetic nervous systems (143). Thus, desires for movement and rest likely operate asymmetrically and are moderately coupled, similarly to the dual system models proposed for energy and fatigue (75), activation and deactivation (144) and positive and negative affect (145). Rest and move could be, at some instances, asymmetric, where individuals do not want to rest, but display also a lack of need to move.

Desires, by definition, have state-like qualities (146), and this appears to be true for desires for movement and rest. Analysis of NOW and WEEK factors of the CRAVE subscales confirmed that these versions were related, but distinctly different. These data suggest that participants were able to differentiate their urge/desires at the present moment versus their typical desires over a much longer period of time. As a state, one might surmise that movement and rest desires vary in patterns similar to state measures of affect (i.e., arousal, pain, energy and fatigue).(49, 129, 130, 136). Indeed, this is what was found. With a maximal treadmill test, desires/wants to move decreased and desires to rest increased, and with an hour-long lecture period, changes were also observed. It is possible that desires/wants for rest and movement may fluctuate diurnally and may further undulate with feedings, sleep, and lifestyle physical activity interspersed throughout the day (144). Future research should aim to understand how desires to move and rest vary across the day with arousal, energy, fatigue, distress and other components of affect. It would be useful to determine what other factors affect movement desires, such as environmental cues, such as lighting (147), music (37, 38), social interactions (148), et cetera (24, 146).

It can be said with near certainty, however, that desires/wants to move and rest fluctuate across time and capture a motivational state.

On the other hand, one might construe that urge and desires for activity and sedentary behaviors are stable and/or may be highly influenced by intra-individual traits. Substantial individual differences have been noted not only in physical activity, exercise and non-exercise activity thermogenesis (NEAT) (149), but also in the acute averse and rewarding effects of physical activity (6, 150). Such a trait would likely possess a strength that is normally distributed throughout the population and expressed most strongly in young age (44). Some children demonstrate more active temperaments, kinetic personalities (i.e., hyperactivity), or a *predilection* for physical activity while others are ostensibly more lethargic (151), traits that may persist into adulthood (152, 153). In a community sample of adults (Study 3), it was found that motivation states trended down (for move) and up (for rest) over a two-year period. As such, more resources are needed to understand how desires might evolve over an extended period of time (e.g., seasonally, over years) and how they can be intervened upon.

Although addressed later in analyses, perhaps the most important hypothesis concerns the very existence of desires/wants to move and rest. It was hypothesized that study respondents would indicate that they perceived desires/wants for movement and rest. In other words, they would not respond with 0 (“none at all”). This is exactly what was found. On the one hand, one might consider this conclusion to be obvious and a trivial matter. However, there is some debate as to whether humans desire movement to a level that it is noticeable. Rosa and colleagues (90) have argued that exercise or physical activity, in themselves, at least from an evolution viewpoint, cannot be primary motivators of behavior. In other words, movement is a by-product, motivated by some other goal, need or extrinsic factor, a means to other ends. Any want of movement would always be overshadowed by desires to rest, be inactive and minimize energy expenditure (91). This line of reasoning holds that humankind evolved a thrifty gene, to spare energy and movement is merely a necessity to avoid harm or to obtain food, shelter and other desired things (54, 123). As modern humans rarely need to move to accomplish these things, the desire to move would likely be minimal. Moreover, one could point out that exercise is fatiguing, painful and possibly feels punishing, thus not reinforcing (54), some individuals clearly do not enjoy exercise or physical activity (154), humans are “hard-wired” to be sedentary (155, 156), and a large portion of the population is, indeed, inactive (91). All of these phenomena likely developed in response to the human need to avoid a negative energy balance.

The counter-arguments indicating the presence of desires/wants to move are equally numerous. First, humans have a strong drive for stimulation (157), and movement may satisfy some of that need. Interestingly, hunter-gather societies engage in frequent cycling of sitting and standing, switching about every 30 minutes, even on highly inactive days (124). Furthermore, as human movement is adaptable and results in greater environmental fitness, it seems likely that it would be wanted at some basic level (123). Even in modern humans, exercise is rewarding for many individuals, similar to food or money (158). The WANT model predicts that desires/wants for movement might even reach levels of strong urges or cravings (17). Such cravings are known to exist, in cases of psychiatric illness (35), neuromuscular disorders (31, 159), or exercise addiction/dependence, in particular (34). Those who “crave activity” likely have rewards systems that activate in response to physical activity and exercise (6, 47). Neurological and genetic underpinnings of desires to move have been identified (see Stults-Kolehmainen for a short discussion) (44, 47, 160). Such pathology is relatively contained to special populations or in response to specific stimuli (37, 38); however, begging the question of whether they are relevant for the general population. This study is one of the first to concretely demonstrate that humans clearly perceive themselves as having desires to move and these are separable from desires to rest.

To understand these processes, future studies should aim to assess movement and rest desires in a variety of populations. The CRAVE was developed in a sample that was ethnically diverse (e.g., in Study 2, participants were 52.5% non-Caucasian and in Study 4, 57.1% non-Caucasian), but the scale should be validated in a variety of different countries, cultures and languages. Furthermore, except for Study 3, the current studies were mostly limited to young adults completing undergraduate education. Since the mid-1800s, it has been submitted that while desire is especially strong for the young and those well-trained, it declines with age and deconditioning (161-164). One might expect that training status, fitness and habituation to physical activity would have an impact on desires for movement. For instance, sedentary and low-active individuals are more likely to experience displeasure during exercise even when it’s low intensity (165). Furthermore, regular exercisers experience greater improvements in mood than non-exercisers (166), and highly active individuals report that enjoyment is the primary motive of exercise (97). Indeed, in the current analysis stage-of-change for exercise predicted wants/desires for movement. However, it would be useful to determine whether those with higher movement desires actually have more energy expenditure (EE) from exercise or non-exercise sources (e.g., occupational activity, active transit, spontaneous physical activity [SPA]) (52, 149) and whether these change over time with chronic training. Movement desires and urges, or what has been termed “appetence”, is magnified with exercise addiction, anorexia, and possibly mania (9, 167). Thus, it is likely, but still speculative, that desires vary over time and across populations, situations and conditions.

Conversely, the desire to move and rest may decrease with a variety of conditions. Movement and rest wants desires likely are altered by conditions like acute and chronic illness (168), depression (psychomotor retardation (169), and distress (psychomotor agitation) (170), which may modify incentive salience for movement (14, 171, 172). Some neuromuscular disorders, like Parkinson’s, are characterized by *apathy* for motor tasks -a near total lack of desire to move (159). As early as 1896, Stedman noted that with nervous and mental disease “the natural conscious *craving* for exercise is lost” (173). Overtraining and exercise burnout have depression-like symptoms and would likely result in altered wants/desires. If such conditions are strong enough, they may even result in dread or aversion for movement as highlighted by the Affect and Health Behavior Framework (AHBF) (15, 20) and further conceptualized by Stults-Kolehmainen et al. (17). Some have argued that the concept of dread for movement is more important as a target for public health initiatives than wants/desires (54). Unfortunately, the current studies were not designed to address the concept of movement avoidance, aversion or dread, and many unknowns exist for these concepts, such as their basis in the brain and neural circuitry (for a review, see (51)). How desires/wants and dread interact should be actively investigated.

Future research should also address how these desires actually motivate to spur behavior (15, 20). Desires vary in magnitude, focal attention and their effect on working memory, which results in differential implications for a wide swath of behaviors (14). Intense desires, manifested as psychophysiological cravings and urges, strongly predict future behavior, such as overeating, binging on alcohol, overconsumption of caffeine, excessive engagement in sexual activity, smoking of cigarettes and other behaviors (174-176). Casper and colleagues documented that the increased urge to move in severely underweight adolescent anorexia nervosa patients was positively associated with exercise intensity and actual exercise over a two-week period (52, 60). As demonstrated by the Dynamical Model of Desire (11), wants that emerge into focal attention in working memory often instigate active pursuit of that desire. Weak, transient desires, on the other hand, may go unnoticed, overshadowed by other desires, and may not result in any changes in behavior (137, 146). However, desires not in focal attention may still motivate behaviors with impelling force through an automatic route of influence. This may be the case for movement, the desire of which is often of a weaker nature, has a smaller demand for attention and exerts less influence on working memory than food or sleep. Nevertheless, it’s possible that desires for energy expenditure may play a key role in influencing movement behaviors and may help to explain compensatory responses to exercise, such as a substantial decrease in activity right after vigorous muscular exertion (9). The CRAVE scale may be utilized to investigate this array of complex processes.

### Conclusions

While theoretical models for cravings/desires for exercise have emerged, the systematic investigation of these constructs is still in its infancy, having been impeded by a lack of instrumentation. We developed a novel measurement tool, the CRAVE, to gauge the want/desire of movement and rest both right now and in the past week. These were tested in 5 samples: 2 large groups of undergraduate students tested cross-sectionally, 2 groups tested multiple times per session and 1 group of community-dwelling adults assessed over a 2-year time period. Exploratory structural equation modelling (ESEM) determined that this instrument has good psychometric properties, distinguishing between desires for movement and rest. It is also sensitive to the state-like nature of the construct. Small changes were observed over a lecture period and large changes were observed with a bout of maximal exercise. Desires to move and rest were associated with perceptions of physical energy, fatigue and tiredness. Consequently, the CRAVE has good construct validity. It also served to test facets of the WANT model (17), which were all supported. Interestingly, movement desires were consistently higher than those for rest. Such a tool may be a valuable resource for researchers interested in psychological wants/desires and how they relate to changes in facets of physical activity and sedentary behaviors in the present moment. It may be particularly useful for those studying exercise addiction / dependence, aging, responses to music (i.e., “groove”), neuromuscular disorders, rehabilitation, and interventions to promote physical activity. Future work should determine how wants/desires vary by environmental and within and between person factors and whether promoting desires for movement and/or resisting desires for rest results in greater movement behavior.

## Supporting information

Supplement to Pre-Print

Crave Scale

## Data Availability

The data and statistical code are available upon request

## Acknowledgements

Marcus Kilpatrick, Ph.D. (University of South Florida) and Justin B. Moore, Ph.D. (Wake Forest School of Medicine) provided thoughtful comments on this project in its early stages. We would also like to acknowledge Regina Casper, M.D., Ph.D., (Stanford School of Medicine), J.D. Hecht, Terry Lu, Ph.D., Asiya Way (University of New Haven) and Samantha Brown (Icahn School of Medicine) for assistance with data management, analysis and interpretation. We also thank numerous graduate students from Teachers College, Columbia University and Southern Connecticut State University, who provided many provoking insights on this project.

