## Supplement to Pre-Print for "Measurement of motivation states for physical activity and sedentary behavior: Development and validation of the CRAVE scale"

### Supplement to Preprint

### Modification Indices and Error Covariances

In the original analyses of the CRAVE scale, modification indices generated by SPSS were utilized to guide model re-specification (1). Error terms were then correlated if a cut-off of 10.0 was detected for the modification index (2). Modelling such correlations based on modification indices has precedence, may result in better model fit, is commonly found in statistical software, and has been used extensively in the past (3-6) (see Gerbing and Anderson for examples (7)). In some cases, the use of modification indices and error term correlations are recommended as a way to detect common method errors (8), which can be highly problematic. Perry (2) evaluated 8 common exercise psychology scales and found that none of them achieved adequate fit unless modification indices were utilized to correlate error terms. Indeed, model specification in CFA has proved difficult across numerous similar exercise psychology measures (2, 9).

Unfortunately, correlating error terms may, a) obfuscate important theoretical constructs and b) may also prevent replication in future studies (7, 9). Their use also appears to be utilized indiscriminately by some researchers just to achieve a better model fit, or perhaps because some researchers have been misinformed on the use of modification indices and error term correlations (as in our case). Due to concerns but also because of continued interest in their use, it has been widely suggested that error term correlations must meet various justification criteria, many of which are still controversial (8, 10-13). Perry and colleagues have noted, "researchers may consider the use of MI to improve model fit" but "do so with caution, and be able to theoretically justify their respecifications" (2). In the preprint describing the original development of the CRAVE scale (14), some of the correlated errors were theoretically justifiable, such as the error correlations between items 13 and 14 ("lay down" and "rest my body"), while others were not as clearly justifiable ("be a couch potato" and "be motionless"). See Figure 1 in the original document.

In Perry's analysis of common exercise psychology scales, most models were significant when examined with exploratory structural equation modelling (ESEM). The authors urge that ESEM be used "from the outset" in exercise psychology research (2) to assist with issues of model fit. This technique was utilized in the revised CRAVE analysis, and correlated error terms were eliminated from models. Such models achieved satisfactory fit, as described in the revised paper.
