## Supplementary material for "Measurement of motivation states for physical activity and sedentary behavior: Development and validation of the CRAVE scale": Crave Scale

**ID________ DATE________ TIME________**

Indicate how much you **WANT** or **DESIRE** to perform the following activities by circling the number along each line between 0 (NOT AT ALL) and 10 (MORE THAN EVER).

Do NOT think about how much you "should" desire each activity.

Think about how you actually want/desire to behave in these ways **at this very moment (i.e., RIGHT NOW)**.

**At this very moment (right now) I want/desire to…**

| | | | |
|---|---|---|---|
| 1) … move my body | NOT AT ALL | 0-----1-----2-----3-----4-----5-----6-----7-----8-----9-----10 | MORE THAN EVER |
| 2) … be physically active | NOT AT ALL | 0-----1-----2-----3-----4-----5-----6-----7-----8-----9-----10 | MORE THAN EVER |
| 3) … do nothing active | NOT AT ALL | 0-----1-----2-----3-----4-----5-----6-----7-----8-----9-----10 | MORE THAN EVER |
| 4) … just sit down | NOT AT ALL | 0-----1-----2-----3-----4-----5-----6-----7-----8-----9-----10 | MORE THAN EVER |
| 5) … burn some calories | NOT AT ALL | 0-----1-----2-----3-----4-----5-----6-----7-----8-----9-----10 | MORE THAN EVER |
| 6) … expend some energy | NOT AT ALL | 0-----1-----2-----3-----4-----5-----6-----7-----8-----9-----10 | MORE THAN EVER |
| 7) … be still | NOT AT ALL | 0-----1-----2-----3-----4-----5-----6-----7-----8-----9-----10 | MORE THAN EVER |
| 8) … be a couch potato | NOT AT ALL | 0-----1-----2-----3-----4-----5-----6-----7-----8-----9-----10 | MORE THAN EVER |
| 9) … exert my muscles | NOT AT ALL | 0-----1-----2-----3-----4-----5-----6-----7-----8-----9-----10 | MORE THAN EVER |
| 10) … be motionless | NOT AT ALL | 0-----1-----2-----3-----4-----5-----6-----7-----8-----9-----10 | MORE THAN EVER |
| 11) … lay down | NOT AT ALL | 0-----1-----2-----3-----4-----5-----6-----7-----8-----9-----10 | MORE THAN EVER |
| 12) … rest my body | NOT AT ALL | 0-----1-----2-----3-----4-----5-----6-----7-----8-----9-----10 | MORE THAN EVER |
| 13) … move around | NOT AT ALL | 0-----1-----2-----3-----4-----5-----6-----7-----8-----9-----10 | MORE THAN EVER |

Scoring: The CRAVE has two subscales: Move and Rest. To calculate *move*, add the ratings from items 1, 2, 6, 9 and 13. To calculate *rest*, sum ratings from items 3, 4, 7, 8, 10. Unscored filler items are 5, 11 and 12.
